# CNVxplorer: a web tool to assist clinical interpretation of CNVs in rare disease patients

**DOI:** 10.1101/2021.03.19.21253806

**Authors:** Francisco Requena, Hamza Hadj Abdallah, Alejandro García, Patrick Nitschké, Sergi Romana, Valérie Malan, Antonio Rausell

## Abstract

Copy Number Variants (CNVs) are an important cause of rare diseases. Array-based Comparative Genomic Hybridization tests yield a ∼12% diagnostic rate, with ∼8% of patients presenting CNVs of unknown significance. CNVs interpretation is particularly challenging on genomic regions outside of those overlapping with previously reported structural variants or disease-associated genes. Recent studies showed that a more comprehensive evaluation of CNV features, leveraging both coding and non-coding impacts can significantly improve diagnostic rates. However, currently available CNV interpretation tools are mostly gene-centric or provide only non-interactive annotations difficult to assess in the clinical practice. Here we present CNVxplorer, a web server suited for the functional assessment of CNVs in a clinical diagnostic setting. CNVxplorer mines a comprehensive set of clinical, genomic, and epigenomic features associated with CNVs. It provides sequence constraint metrics, impact on regulatory elements and topologically associating domains, as well as expression patterns. Analyses offered cover (a) agreement with patient phenotypes; (b) visualizations of associations among genes, regulatory elements and transcription factors; (c) enrichment on functional and pathway annotations; and (d) co-occurrence of terms across PubMed publications related to the query CNVs. A flexible evaluation workflow allows dynamic re-interrogation in clinical sessions. CNVxplorer is publicly available at http://cnvxplorer.com

## INTRODUCTION

To date, more than 5,000 Mendelian phenotypes have been clinically recognized. Thus, differential diagnosis of such heterogeneity based on human recognition of clinical symptoms is challenging even for the most experienced medical geneticists. Moreover, around 50% of all known Mendelian diseases still lack the identification of the causal gene or variant (1). In addition to SNVs and indel variants, Structural Variants (SVs) involving large unbalanced genomic rearrangements such as deletions and duplications (Copy Number Variants, CNVs) are a major cause of Mendelian diseases (2, 3). Proper identification of CNVs can help further increase diagnostic rates (4). For instance, recent work reported pathogenic CNVs in 9% of patients presenting inherited bone marrow failure syndrome (IBMFS) (5), in 10,5% of individuals with neurodevelopmental disorders (6), and 8% of the pathogenic Primary Immune Deficiency (PID) variants found in Stray-Pedersen et al. (7) were CNVs.

In a clinical setting, array comparative genomic hybridization (aCGH) performed on DNA from whole blood cells is currently the primary genetic test to detect CNVs (resolution ∼150-300 Kb). The French network of cytogeneticists and molecular geneticists, Achro-Puce (8) reported in 2019 a total of 16,993 aCGH tests prescribed to pediatric patients presenting rare disease phenotypes. Among them, geneticists classified 11,8% as bearing pathogenic or likely pathogenic CNVs. However, a significant fraction of patients (8,2%, 1,393 / year) presented CNVs labeled as variants of unknown significance (VUS), revealing current clinical interpretation limitations. Challenges in CNV clinical evaluation arise from the difficulty to interpret functional consequences on genomic elements other than those overlapping with disease-associated genes or with previously reported pathogenic CNVs.

Recent studies have shown that a more comprehensive evaluation of clinical, genomic, and epigenomic features, leveraging both coding and regulatory impact, can help improving diagnostic rates (9, 10). However, currently available CNV annotation tools are mostly based on the assessment of the protein-coding genes targeted by the CNVs under evaluation (e.g., SG-ADVISER CNV (11) and cnvScan (12)) or aggregate static annotations from reference databases (AnnotSV, (13)) without providing further filtering and analysis tools. In addition, features of interest often involve heterogeneous high-dimensional *omics* data, scattered across multiple databases, often difficult to access and interpret from a clinical perspective. In practice, the evaluation task may be error-prone, time-consuming, and lowly reproducible across geneticists. Thus, there is a need for interactive computational interfaces and pathogenicity scores facilitating the accuracy and reproducibility of CNV assessment.

To address these challenges, we developed CNVxplorer, a user-friendly web application that facilitates the clinical interpretation of CNVs, irrespective of the technology used to identify them. In addition to the overlap with reference sets of pathogenic and benign CNVs, SNVs, and disease genes, CNVxplorer’s output provides a comprehensive set of features associated with the query CNVs, including (i) sequence conservation scores both across species and within humans (10); (ii) gene dosage sensitivity estimates (e.g., haploinsufficiency or triplosensitivity); (iii) overlap with non-coding genes (lncRNAs and miRNAs), enhancers, transcription factors, and Topologically Associated Domains, (TADs) (14, 15). More distal genes regulated in cis or trans by the impacted regulatory regions can also be incorporated into the analysis (9).

CNVxplorer downstream analyses include assessing the phenotypic similarity between the associated genomic elements and the patient clinical signs coded as Human Phenotype Ontology (HPO) (16) and graph visualizations of gene-gene associations mediated by regulatory elements and transcription factors. The cumulative effect of the previous aspects on biological processes and on molecular pathways (17) is evaluated through gene-set enrichment analyses. Finally, PubMed publications related to the query CNVs and their underlying network of term co-occurrence are presented. CNVxplorer is designed as an interactive tool allowing geneticists to update results “on-the-fly” after modifying filtering thresholds, parameter settings, and the set of retained genomic elements for downstream evaluation. CNVxplorer is publicly available at http://cnvxplorer.com. Detailed tutorials, comprehensive documentation, and a Frequently Asked Questions section are provided. In addition, a stand-alone open-source R implementation with a *shiny* interface is offered, allowing its deployment as a private server through a *Docker* image without external dependencies. Source code and instructions to locally deploy the application are available at https://github.com/RausellLab/cnvxplorer.

## MATERIAL AND METHODS

### Genetic variants datasets

CNV syndromes were obtained from DECIPHER (version 2020-01-19, (18)) and ClinGen (19) databases. Pathogenic and likely pathogenic CNVs were obtained from DECIPHER (version 2020-01-19, (18)) after filtering out CNVs of length < 50 base pairs. CNVs identified in the general population were obtained from the DECIPHER control set as well as from the DGV database (version 2020-02-25, (20)) and gnomAD (controls-only dataset, version 2.1, (21)). DGV CNVs were restricted to those labeled as “deletions”, “duplications”, “loss”, and “gain”, while gnomAD (21) CNVs were reduced to those with a filter equal to PASS and SVTYPE equivalent to ‘DEL’ or ‘DUP’. Pathogenic and likely pathogenic variants were further extracted from ClinVar (version 2021-02-21, (22)). Trait-associated Single Nucleotide Polymorphisms were obtained from the GWAS Catalog (version 1.0, (23)). GWAS variant coordinates were converted from hg38 to hg19 using UCSC liftOver (24) De novo variants were obtained from denovo-db (both Simons Simplex Collection (SSC) and non-SSC samples, version 1.6.1, (25)) considering only loss-of-function variants (“frameshift”, “splice-donor”, “stop-gained”, “start-lost”, “splice-acceptor”) and excluding variants from control samples. Segmental duplications and low-mappability regions were extracted from the ENCODE Blacklist (26).

### Gene-level annotations

A list of 18,792 protein-coding gene symbols was retrieved from HUGO Gene Nomenclature Committee (HGNC, version 2021-03-03) (27). Transcript boundaries were obtained with BioMart (28) from Ensembl Gene (version 103, (29)) based on the reference human genome GRCh37.p13. Disease gene lists were collected from five sources: Online Mendelian Inheritance in Man (OMIM) database (version 2021-03-04, (30)), Orphanet (version 2021-03-04, (31)), DECIPHER (version 2020-01-19, (18)), Genomics England PanelAPP (2021-03-04, (32)), and ClinGen (version 2021-03-03, (19)). Following Caron et al. (33) OMIM genes were restricted to monogenic Mendelian disease genes by filtering out genes associated with phenotype descriptions flagged as “somatic” or “complex” and with a supporting evidence level of 3 (i.e., the molecular basis of the disorder is known). DECIPHER disease genes were restricted to those labeled as “confirmed”. Clingen genes were restricted to (i) those with a score equal to 3 (i.e., sufficient evidence of dosage sensitivity) in the case of haploinsufficiency genes, and (ii) those with a score equal or higher than 1 (i.e., little evidence of dosage sensitivity) in case of triplosensitivity genes. Clingen genes with a score equal to 40 (i.e., dosage sensitivity unlikely) were excluded. Associations between genes, OMIM and ORPHANET disease identifiers were downloaded from http://purl.obolibrary.org/obo/hp/hpoa/phenotype.hpoa.

Gene-level scores for sequence constraint in humans included the Residual Variation Intolerance Score (RVIS, version 3, (34)), the gene probability of LoF intolerance (pLI, version 2.1.1, (35)), the Constrained Coding Region score (CCRs, (36) and the Gene Damage Index (GDI, a metric that was designed to assess the mutational damage amassed by a gene in the general population (37). Interspecies conservation was evaluated through the estimated proportion of nonlethal nonsynonymous mutations, which was obtained with the selection inference using a Poisson random effects model (SnIPRE), by comparing polymorphism within humans and divergence between humans and chimpanzee at synonymous and nonsynonymous sites (38). Non-coding versions of RVIS (ncRVIS) and the Genomic Evolutionary Rate Profiling rejected substitutions (ncGERP), based on proximal regulatory regions (5’UTR, 3’UTR, 250bp upstream of TSS), were further considered (39). In addition, the gene predicted probability of haploinsuficency score was retrieved (HI, (40). Gene essentiality labels were obtained from the FUSIL classification (41). Imprinted genes were obtained from publicly available imprinting gene database (http://www.geneimprint.com/) and ohnolog genes from the OHNOLOGS database (version 2, (42)) considering only pairs labeled as “strict” (highly reliable). Ohnologs are gene duplicates originated and retained from the two rounds of whole-genome duplication (WGD) that occurred in early vertebrates ∼500 million years ago (43).

Gene expression data for 54 human tissues was obtained from the Genotype-Tissue Expression Project (GTEx; version 8, (44)). Protein expression levels for 32 human tissues was obtained from the Human Protein Atlas (HPA; version 20.1, (45); https://www.proteinatlas.org/download/normal_tissue.tsv.zip). The HPA project provides a curated reliability score based on the results’ reproducibility and data availability in other databases. Thus, only protein expression levels labeled with “Enhanced” or “Supported” reliability were considered, while those labeled as “Not detected” are not reported in the application. The list of protein-protein interactions was obtained from the STRING database (version 11, (46)). STRING provides a set of experimental and predicted genes and protein interactions extracted from diverse sources. Sources considered in CNVxplorer include physical protein-protein interactions determined through biochemical assays,co-expression data,co-citation in abstracts from scientific literature, evolutionary gene-fusion events, and co-presence in the same genomic neighborhood across species. Only high confidence STRING interactions (> 700 score) were retained.

### Regulatory elements and associated genes

Enhancer regions and their associations with target-genes were obtained from the GeneHancer database (version 4.11, (47)). Predicted enhancers and enhancer-gene associations with a score < 1 were filtered out, resulting in a total of 107,168 enhancer-target pairs finally retained. Topologically Associating Domains (TAD) genomic coordinates were obtained from (48), corresponding to H1 human embryonic stem cells (hESCs) and generated using a bin size of 40 kilobases and a window size of 2 Mb. Long non-coding RNAs (lncRNAs) and their associated genes -characterized by low throughput experiments-were obtained from LncRNA2Target (version 2, (49)). Transcription factors and their associated genes were retrieved from TRRUST (version 2, (50)). miRNAs coordinates were retrieved from miRbase (51) and miRNA-gene associations with strong experimental evidence were obtained from miRTarBase (version 8, (52)). Conservation estimates of regulatory elements across species were calculated as the mean PhastCons score per base pair using 99 vertebrate species, 45 placental mammals, and 45 primates multi-species alignments from UCSC. Enhancers and miRNAs coordinates were converted from hg38 to hg19 using UCSC liftOver (24). Segmental duplications and low-mappability regions were extracted from the ENCODE Blacklist ((26), version 2).

### Phenotypic annotations and clinical similarity assessment

Phenotypic terms associated with human genes and diseases were retrieved from the Human Phenotype Ontology (HPO, version 2021-02-09, (16)). Suggestions of additional HPO terms were automatically generated based on the descendent terms of the HPO input list. When multiple descendent terms were found, those with the highest Information Content (IC) were retrieved. Calculations of the phenotypic similarity between two sets *Q* and *D* of *t* HPO terms, was done using symmetric Resnik’s measure and aggregated following the Best Match Average (BMA) method (53):

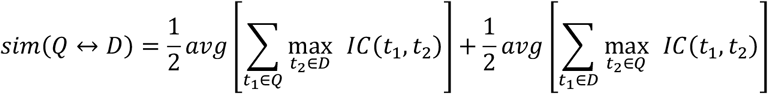

Anatomical entities are defined as the set of HPO terms directly descendending from the parent term *phenotypic abnormality* (HP:0000118), e.g., abnormality of limbs, abnormality of the eye, etc. Anatomical entities were identified from HPO term annotations (and their parent terms) of genes and diseases. In the case of genes, HPO terms correspond to their disease-associated annotation. Automatic text annotation of phenotypic entities is performed through the SciGraph API (54) using the R package bioloupe (version 0.0.0.9000, (55)). Mouse/Human orthology relations and phenotype data resulting from knockout experiments in mice were retrieved from Mouse Genome Informatics (MGI; version 2021-03-01; (56)). Description of mouse phenotype terms was obtained from the Mammalian Phenotype Ontology (MPO; version 2021-01-12; (57)).

### Biomedical literature

PubMed articles were retrieved from the NCBI Entrez system using the R package rentrez (version 1.2.2, (58)). For each query CNV, the associated cytoband(s) are obtained, and two PubMed queries with the following structure are performed, respectively: Query 1 [*((chromosome identifier + cytoband) OR chromosome-cytoband) AND (deletion OR microdeletion) AND Homo sapiens*]; Query 2 [*((chromosome identifier + cytoband) OR chromosome-cytoband) AND (duplication OR microduplication) AND Homo sapiens*]. For those CNV(s) mapping different cytobands and/or chromosomes, CNVxplorer concatenates the queries for each cytoband with an “OR” separator. The option “Filter articles associated with OMIM entries” is performed using the R package rentrez and searches for OMIM gene and disease records associated with each Pubmed entry that the NCBI previously indexed. Biomedical co-occurrence networks were generated based on the number of paired presence of keywords in article titles and abstracts. A network plot is generated using the R package ggraph (version 2.0.0). Automatic text annotation of genetic, mutation, phenotypic and chemical/drug entities in titles and abstract was performed with PubTator Central API (59) using the R package bioloupe (version 0.0.0.9000).

### Functional analysis

Functional and pathway enrichment analysis were implemented with the R/Bioconductor packages ClusterProfiler (version 3.14.3; (60)), and reactomePA (version 1.30.0, (61)). KEGG (62) and REACTOME (63) pathway annotation databases were used. Disease ontology enrichment analysis and visualization are implemented with the R/Bioconductor package DOSE (version 3.12.0, (64)). Tissue enrichment analysis is implemented with the R/Bioconductor TissueEnrich package (version 1.6.0, (65)).

### Implementation of CNVxplorer web application and docker image

CNVxplorer was developed in R (version 3.6.0, (66)). The interactive web application was implemented with the Shiny R package (http://shiny.rstudio.com). This framework allows the fast implementation of R code in an easy-to-use frontend. The interface offers a reactive environment where the user can interact with the data (e.g., add/remove input) and the results are instantly updated accordingly. CNVxplorer was additionally implemented as a Docker image (https://www.docker.com/). This facilitates the installation of the application in a private server mode without installing additional external dependencies or prerequisites. All plots and tables presented in CNVxplorer were rendered with ggplot2 and DT packages, respectively. Interactive network visualization was constructed with network3D R package. The visual interface was enhanced with the tablerDash package. Details about the database and R package versions used are available on the CNVxplorer website (***Documentation*** tab, section ***Input data and software***).

## RESULTS

### Workflow

CNVxplorer allows simultaneous analysis of single or multiple CNVs, irrespective of the technology used to identify them. It requires as input either a single genomic interval, a cytoband, or a .tsv file with multiple entries. Optionally, the patient phenotypes can be further reported in downstream analyses coded as HPO terms (**Methods**). Once the query is submitted, the application displays the results across nine tabs (**Figure 1**), namely: (1) overview, (2) clinical genetic evidence, (3) regulatory regions, (4) protein interaction, (5) phenotypic analysis, (6) KO mouse phenotypes, (7) biomedical literature, (8) tissue specificity, and (9) functional enrichment analysis. The information and analyses provided across these tabs is presented in detail in the following subsections.

**Figure 1.**
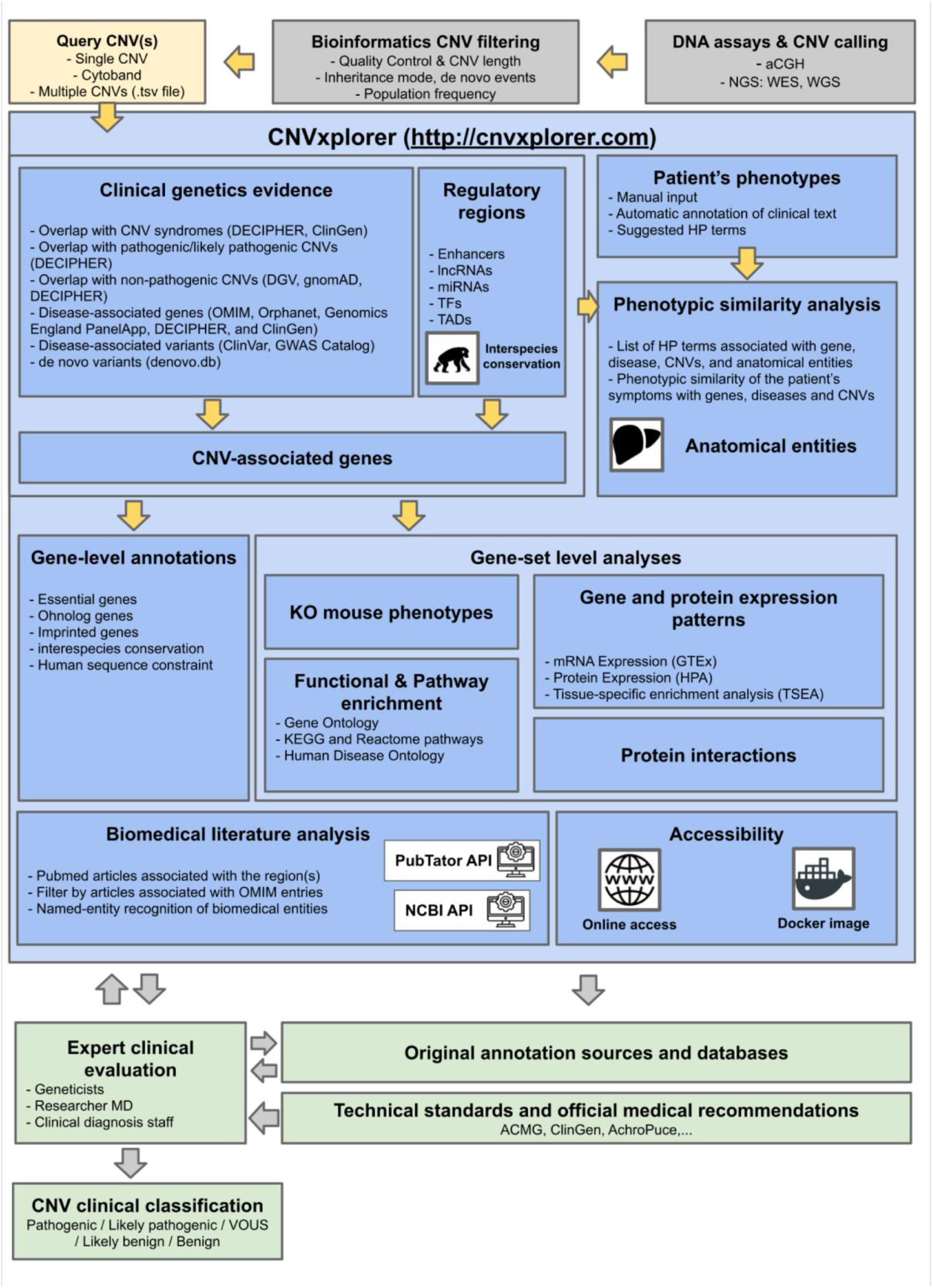
CNVxplorer overview and integration within the clinical diagnosis workflow. The figure represents the annotation process and analyses offered by CNVxplorer together with its integration in the clinical workflow for genetic diagnosis of rare disease patients. CNVxplorer provides analyses tools for CNV interpretation the irrespective of the technology used to identify them. Thus, CNV calling together with prior bioinformatics filtering needs to be done upstream the use of the application. CNVxplorer offers a dynamic workflow for the investigation of a comprehensive set of genomic, epigenomic and clinical features together with diverse gene-set enrichment analyses and the automatic exploration of the biomedical literature. The application is interactive allowing clinical experts to re-evaluate the analysis process in an iterative way. Finally, it corresponds to the geneticist, researcher MD or clinical diagnosis staff to provide CNV clinical classificatios following technical standards and medical recommendations and after further checking the original annotation sources for in-depth details. Abbreviations: CNV: Copy Number Variants; aCGH: array comparative genomic hybridization; NGS: Next Generation Sequencing; WES:Whole Exome Sequencing; WGS: Whole Genome Sequencing; lncRNAs: long non-coding RNAs; miRNA: micro RNAs; TF: Transcription Factors; TADs: Topologically Associating Domains; ACMG : American College of Medical Genetics and Genomics; ClinGen : Clinical Genome Resource; Achro-puce: network of French cytogeneticists; DECIPHER: Database of Chromosomal Imbalance and Phenotype in Humans Using Ensembl Resources; OMIM: Online Mendelian Inheritance in Man; HP term: Human Phenotype term; KO: Knock-out; MGI: Mouse Genome Informatics; GO: Gene Ontology; GTEx: Genotype-Tissue Expression project; HPA: Human Protein Atlas; TSEA: Tissue-specific gene enrichment analysis; NCBI: National Center for Biotechnology Information.

***Tab 1: Overview***. This section provides a quick overview of the major clinically relevant associated with the query CNV, summarizing the results presented in full detail in the next tabs. Thus, it provides summaries of the CNV’s length, gene content, overlap with reference CNV databases, as well as with disease-associated genes and single nucleotide variants together with the number of disrupted regulatory elements. In addition, the number of orthologous mouse genes associated with mortality, aging phenotype or embryonic lethality is reported. A popup warning is displayed if any input CNVs overlap with a segmental duplication region or a low-mappability region (**Methods**). Complementary information includes the number of PubMed articles where the CNV associated cytobands are mentioned together with the keywords *duplication* or *deletion*. Finally, the size of each of the query CNVs is represented together with the size distribution of CNVs found on the reference databases (i.e., gnomAD, DGV, Decipher Control set, and Decipher pathogenic set; **Methods**).

***Tab 2: Clinical genetic evidence***. This section provides an exhaustive report of disease-associated CNVs, genes, and SNVs that partially or totally overlap with the query CNV(s). First, a list of CNVs previously reported in reference pathogenic and non-pathogenic CNV sets is provided, covering (i) CNVs associated with a phenotype, including CNV syndromes as well as pathogenic or likely pathogenic CNVs, as reported in the CinGen or Decipher pathogenic sets (**Methods**), and (ii) CNVs classified as benign, likely benign or present in the general population, as reported in Decipher control, DGV and gnomAD datasets (**Methods**). Provided details include genomic coordinates, percentage of overlap with query CNV(s), associated phenotypes, or frequency in the reference sets. Information is displayed as tables allowing to search, sort, and filter by column values. Second, a table shows all protein-coding genes mapping within the CNV genomic coordinates, indicating whether they are disease-associated or essential genes. A suplemmentary panel summarizes the number of disease-associated genes found across the different interrogated sources (OMIM, Orphanet, DECIPHER developmental disease genes, Genomics England Panels and ClinGen; **Methods**). CNVxplorer additionally integrates gene-level annotations relevant for clinical interpretation, such as their ohnolog or imprinted nature, along with the expressed allele (maternal or paternal) (**Methods**). A complete set of gene-level computational pathogenicity scores is reported, including metrics of sequence constraint in humans (pLI, CCRs, GDI, RVIS, and non-coding RVIS), inter-species conservation (ncGERP), variation within and between species (SnIPRE), and haploinsufficient genes prediction from machine-learning approaches (HI index). To ease clinical interpretation, previous scores are displayed as genome-wide rank-percentiles, ranging from 0 (non-pathogenic) to 100 (pathogenic). Finally, two additional tables are provided showing (i) disease-associated variants from ClinVar and the GWAS catalog, and (ii) *de novo* variants identified in trio studies using whole-exome and whole-genome sequencing (**Methods**).

***Tab 3: Regulatory regions***. CNVxplorer displays a comprehensive list of regulatory elements mapping within the CNV(s) coordinates, displayed across five tables corresponding to enhancers, miRNAs, lncRNAs, genes encoding Transcription Factors (TFs), and Topologically Associating Domains (TADs), respectively (**Methods**). Details provided include genomic coordinates and the target gene associated with each of the regulatory elements found. Thus, the user has the option for each of the five tables to include those target genes in the input list used by the downstream gene-based analyses of the application, even if these genes do not directly overlap with the query CNV(s). Those target-genes not mapping with the CNV(s) are collectively displayed at the top of the tab, where several tables with gene-level annotations analogous to the ones described in ***Tab 2*** allow to assess their clinical and functional significance. Complementary information for regulatory elements includes the action mode of transcription factors (activation or repression), disease-associations of long non-coding RNAs, and enhancer conservation scores based on vertebrate, mammal, and primate genome alignments (**Methods**). Thus, upon selection of a given enhancer, its conservation is plotted in 3 panels representing the background distribution of conservation for all human enhancers across vertebrates, mammals, and primates, respectively. Such conservation scores can be regarded as proxies of their functional redundancy and tolerance to loss of function mutations (18).

***Tab 4: Phenotypic analysis***. This tab aims to evaluate the phenotypic agreement between the genomic elements targeted by the query CNV(s) and the clinical signs observed in the patient under evaluation. For this, CNVxplorer first displays the top 10 most frequent HPO terms and the top 10 most frequent anatomical entities to which the CNV-target genes (as retained in **Tab 2** and **Tab 3**) are associated with (**Methods**). Second, CNVxplorer requests the user to provide a list of Human Phenotype Ontology (HPO) terms describing the patient’s clinical status. The user can provide such input in two ways: either by manually selecting HPO terms from a reference table that supports text search, or ii) by providing a free text description of the patient’s signs, which is then processed by the application to automatically extract HPO terms (**Methods**). The previous array of the patient’s HPO terms is then compared against the HPO terms associated with three types of instances:.(i) the genes related to the query CNV(s), (ii) the diseases associated with the previous gene list (which can be filtered by inheritance mode, e.g., autosomal dominant, recessive, etc.), (iii) the pathogenic CNVs overlapping with the query CNV(s) (as presented in **Tab2)** (**Methods**). As an output, CNVxplorer provides the clinical similarity between the patient’s phenotypes and each of the previous three instances, independently evaluated. Thus, rankings by phenotypic similarity are then plotted for genes and their associated diseases as well as for the pathogenic CNVs associated with the genomic intervals of the query CNV(s). Phenotypic similarity rankings may point to the genomic regions within the query CNV(s) most likely to be responsible for the observed patient’s phenotypes. The application instantly reacts to any change in the HPO input list or free text content by automatically updating the phenotypic similarity scores. Furthermore, upon selection of a specific gene or disease, the application displays a bar plot representing the distribution across anatomical entities of their associated HPO terms and compares them to those of the patient’s HPO terms.

**Tabs 5 - 8. Gene-set level analysis**. CNVxplorer offers here diverse gene-set based analyses for genes mapping within the genomic intervals of the query CNV(s) together (or not) with genes associated with the affected regulatory elements (according to the user’s settings in **Tab 3**). These analyses are:

**Tab 5: KO mouse phenotypes**. The tab presents a histogram of the phenotypic consequences observed in knockout and knock-down mouse experiments for mouse orthologs of the genes under evaluation, collectively considered (**Methods**). Results are displayed separately according to whether the genes map within the query CNV(s) boundaries or outside them (i.e., indirect implication though a targeted regulatory element).

**Tab 6: Functional enrichment analysis**. CNVxplorer provides gene-set enrichment analyses using (i) Gene Ontology (GO) terms (performed by each GO ontology: biological process, molecular function, and cellular compartment), (ii) reference pathway databases such Reactome and KEGG, and (iii) Human Disease Ontology (HDO) terms (**Methods**). Here, the list of functional terms and pathways that are statistically overrepresented in CNV-associated genes in reported together with their hypergeometic test p-value, corrected by Benjamini-Hochberg False Discovery Rate. Results can be displayed in tables or bar plots, and the p-value threshold can be modified according to the user’s preference.

**Tab 7. Tissue expression patterns**. This tab displays gene expression patterns across 54 tissues from the GTEX project and protein expression levels across 32 tissues from the Human Protein Atlas (Methods). Gene expression levels are represented in Transcripts Per Million (TPM) units. Protein expression levels, in contrast, are indicated in a qualitative way (low, medium, and high), as provided by the original sources. A tissue-specific gene-set enrichment analysis can be performed against GTEx tissues.

***Tab 8: Protein interaction network***. This section offers a quick visualization of the relationship among the genes under evaluation, which is represented in the form of a network. In such network, nodes represent genes/proteins are links among them represent a relation or interaction among them. For this, CNVxplorer mines high-confident protein-protein interactions reported in the STRING database ((46), **Methods**). CNVxplorer provides an interactive graphical interface, allowing users to zoom in/out and drag nodes from specific network regions. To ease visualization, gene-centered representations with 1-neighbor nodes are also offered. Finally, a bar plot representing the number of interactors per gene is displayed.

***Tab 9: Biomedical literature***. CNVxplorer eases the exploration of biomedical literature associated with the query CNV(s). First, a comprehensive search across all fields of Pubmed articles is performed (**Methods**), and articles found are displayed as an interactive table allowing: (i) to filter by event type (deletion or duplication) and whether the article is associated with an OMIM entry, (ii) to sort by column content, e.g., on the number of citations or on the year of publication. By selecting a given PubMed entry on the table, its abstract is automatically displayed, and genetic, mutation, phenotypic, and chemical/drug entities are automatically highlighted therein (**Methods**). Additionally, a co-occurrence network of keywords across the retrieved PubMed entries is automatically generated. Here, words are represented as nodes, their co-occurrence across articles is represented as links, and the link thickness is proportional to the number of articles where words co-occurre (**Supplementary Figure 1**). Such netwok provides an graphical overview of the terms’ relevance and their associations.

### Examples of prototypical analysis workflows provided by CNVxplorer in a clinical diagnosis setting

To illustrate the potential of CNVxplorer to help in the interpretation of variants in a clinical setting, we propose two different case studies of CNVs identified in patients presenting congenital abnormalities:

### Case study 1 – Fine mapping of causal genomic elements within a pathogenic CNV

Waespe et al. (67) identified CNVs associated with inherited bone marrow failure syndromes (IBMFS). This group of syndromes is genetically heterogeneous and characterized by hematologic complications. Here we investigate a heterozygous deletion of ∼11.3Mb in chr3(GRCh37):186550246–197837050, reported by the authors in a patient with Diamond–Blackfan anemia. CNVxplorer results for this genomic interval showed a 100% overlap with a CNV syndrome (3q29 microdeletion syndrome) reported both in DECIPHER and ClinGen (***Overview*** and ***Clinical genetics evidence*** tabs). In addition, after restricting to heterozygous and deletion events, a total of 16 pathogenic CNVs from DECIPHER appeared as totally overlapped by the query CNV, which supports the pathogenicity of the variant (***Clinical genetics evidence*** tab). However, a more challenging question is whether a fine mapping of the causal genomic element can be proposed. Here, CNVxplorer found 71 protein-coding genes mapping within the CNV genomic intervals, including 22 disease-associated genes (***Clinical genetics evidence*** tab). To narrow down the number of plausible causal gene(s). First, the patient’s clinical status can be leveraged by CNVxplorer to identify the target genes with the highest phenotypic similarity to the patient’s symptoms (***Phenotypic similarity*** tab). Thus, by indicating the HP term *congenital hypoplastic anemia* (HP:0004810) and after filtering the diseases with autosomal dominant inheritance, the genes RPL35A and LPP appeared as the ones with the highest clinical similarity to the patient’s phenotype. RPL35A is indeed associated with Diamond–Blackfan anemia, while LPP is linked to acute myeloid leukemia. Consistent with this finding, four disease-associated variants from ClinVar are reported as associated with the gene RPL35A and Diamond-Blackfan anemia (***Clinical genetics evidence*** tab). Finally, CNVXplorer searches across PubMed (***Biomedical literature*** tab), after filtering by “deletion” event and by the text “Diamond-Blackfan anemia”, retrieved the PubMed entry PMID: 28432740, further supporting the causal role of RPL35A in the patient’s syndrome.

### Case study 2 – Clinical interpretation of CNVs through their impact in non-coding regulatory regions

Flöttmann et al. (9) reported non-coding CNVs identified in patients with congenital limb malformations. We assess here a deletion of ∼440-kb in chr2(GRCh37):176065894-176504173, reported by the authors in a patient with brachydactyly type E (BDE, HP:0005863). CNVxplorer results for this genomic interval showed a minor overlap (<11.59%) with 16 pathogenic/likely pathogenic CNV deletions from DECIPHER (***Overview and Clinical genetics evidence*** tabs). In turn, a low overlap (<14,52%) was found as well with non-pathogenic CNV deletions from Decipher (n=4), DGV (n=13) and gnomAD (n=16). However, no protein-coding genes were found within the genomic intervals of the query CNV and, while 11 disease-associated SNPs from GWAS studies were found, none of them related to the patient’s clinical signs (***Clinical genetics evidence*** tab). Notwithstanding, an inspection of the regulatory elements mapping within the CNV targeted region showed the presence of an enhancer region in chr2:176469730-176471596. This enhancer is associated with HOXD13 and ATP5MC3 genes, which map outside the boundaries of the query CNV (***Regulatory regions*** tab). The corresponding CNVxplorer panels show that the enhancer region is highly conserved across vertebrates, mammals, and primates, suggesting functional relevance. Further inspection of CNVxplorer results showed that the query CNV partially overlapped with a TAD spanning chr2: 176200000-176960000, thus disrupting its left boundary. Such TAD contains EVX2 and LNPK genes, in addition to HOXD13. By clicking on the “add target genes to the analysis” button both in the enhancer and in the TAD tables, CNVxplorer reacts by displaying their gene features. Here both HOXD13 and LNPK are reported as a disease-associated gene (Regulatory regions tab). Both genes are in turn reported as human ohnologs and, while HOXD13 is indicated as an haploinsufficient gene, LNPK presents high conservation levels of its non-coding regions (ncGERP and ncRVIS than 95 rank percentile). The ***KO mouse phenotypes*** tab shows that knockout/ knock-down mouse experiments lead to a limbs/digits/tail phenotype for the mouse orthologs of HOXD13, EVX2, and LNPK. However, a deeper inspection of the associated MGI links showed that, while such phenotype was observed for a targeted disruption of HOXD13 (i.e., not involving other genes), in the case of LNPK and EVX2, the limbs/digits/tail phenotype associated to inversions involved a large number of genes. The inspection of human phenotypes associated with the two disease-associated genes found, i.e., HOXD13 and LNPK (***Phenotypic analyses*** tab), showed that the brachydactyly type E had been indeed previously associated with HOXD13, further supporting the causal role of the query CNV in the patient’s phenotype. Finally, the **Biomedical literature tab** automatically retreived four PubMed articles citing the entity “HOXD13”, and three citing the entity “syndactyly”, an HPO term related to brachydactyly type E. Accordingly, the entities co-occurrence network obtained further reflected the association of the gene HOXD13 with terms such syndactyly, limb malformations and haploinsufficiency (**Supplementary Figure S1**).

## DISCUSSION

CNVxplorer was conceived as an interactive webserver application to improve the functional assessment of CNVs, with a major focus in genetic diagnosis of rare disease patients. To that aim, the application offers a dynamic exploration of a large set of genomic, epigenomic and clinical CNV features that have proven to be relevant for clinical interpretation ((68, 69)). The analysis tools and workflows implemented ease the exploration of major paradigms in current cytogenetics, i.e. : (i) the consideration of regulatory elements, including 3D chromosome organization, and their associated genes, which may eventually map distal regions outside the CNV boundaries; (ii) the investigation of putative oligogenic effects that could translate into a combined impact on specific biological processes, pathways or tissues, assessed through gene-set enrichment analyses; and (iii) the evaluation of the joint impact that multiple structural events, together with short indels, may have on a single individual genome, which CNVxplorer approaches through the assessment of their combined consequences, instead of providing independent annotations for each CNV event.

Rather than a passive reader of CNV annotations or pathogenicity scores, the user of CNVxplorer (e.g. a clinical geneticist or researcher MD) becomes actively involved in the CNV evaluation process. Full control is given on critical aspects such as the reference CNVs partially overlapped by the query CNVs, the regulatory elements retained, the gene-level filters or score thresholds applied, or the characteristics of the enrichment tests performed, among many others. Several settings can thus be easily explored, and their impact in downstream clinical interpretation instantly evaluated. The reactivity of the application allows immediate discussion during clinical diagnosis sessions. Collegial interpretation is particularly relevant in the assessment of the patient’s clinical signs and how well they fit with the affected genomic elements. CNVxplorer provides thus an extensive toolkit for phenotypic similarity assessment based on HPO terms associated to the CNVs, genes and diseases retrieved, together with automatic text processing tools of the associated biomedical literature.

CNVxplorer offers a rich compendium of annotations and analyses, yet it does not generate as output an automatic classification, neither quantitative nor qualitative, of the pathogenicity potential of the query CNV(s). Such classification is currently left to the expert interpretation of clinical geneticists, who then can proceed according to the official guidelines and technical standards of reference medical associations, such as the joint consensus recommendation of the American College of Medical Genetics and Genomics (ACMG) and the Clinical Genome Resource (ClinGen), in the US ((70)), or the recommendations for CNV clinical interpretation of the Achro-puce network of French cytogeneticists ((71)) (**Figure 1**). Thus, the added value of CNVxplorer in diagnosis accuracy, reproducibility and time efficiency can only be further evaluated through a multi-site retrospective study on unpublished sets of pathogenic and bening CNV variants.

The automatic pathogenicity scoring of CNVs will constitute, nonetheless, a main next step in future CNVxplorer developments. This is now a pressing need, considering the increasing adoption of Whole Genome Sequencing (WGS) as a clinical diagnostic tool, which allows the identification of high numbers of CNVs of small size (72–74)). The success rate in the clinical evaluation of CNVs from WGS would largely benefit from Artificial Intelligence techniques able to narrow down the list of candidate variants for expert annotation. Unbiased training of classifiers, however, remains challenged by the historical ascertainment bias towards pathogenic CNVs of large-size and mostly targeting protein-coding regions (18). In this context, comprehensive CNV analysis tools such CNVxplorer may contribute to accelerate the identification of pathogenic and likely pathogenic CNVs targeting shorter non-coding regions. Such variants can then constitute a reliable corpus for downstream machine-learning developments.

## Data Availability

CNVxplorer is publicly available at http://cnvxplorer.com. Detailed tutorials, comprehensive documentation, and a Frequently Asked Questions section are provided. The tool is free and open to all users and there is no login requirement. CNVxplorer code and instructions to deploy a local Docker image are available at https://github.com/RausellLab/cnvxplorer, under a GNU General Public License v3.0.

https://github.com/RausellLab/cnvxplorer

## ACKNOWLEDGEMENT

This study makes use of data generated by the DECIPHER community. A full list of centres who contributed to the generation of the data is available from https://decipher.sanger.ac.uk/about/stats and via email from. Funding for the DECIPHER project was provided by Wellcome.

## FIGURES and FIGURE LEGENDS

**Supplementary Figure 1.**
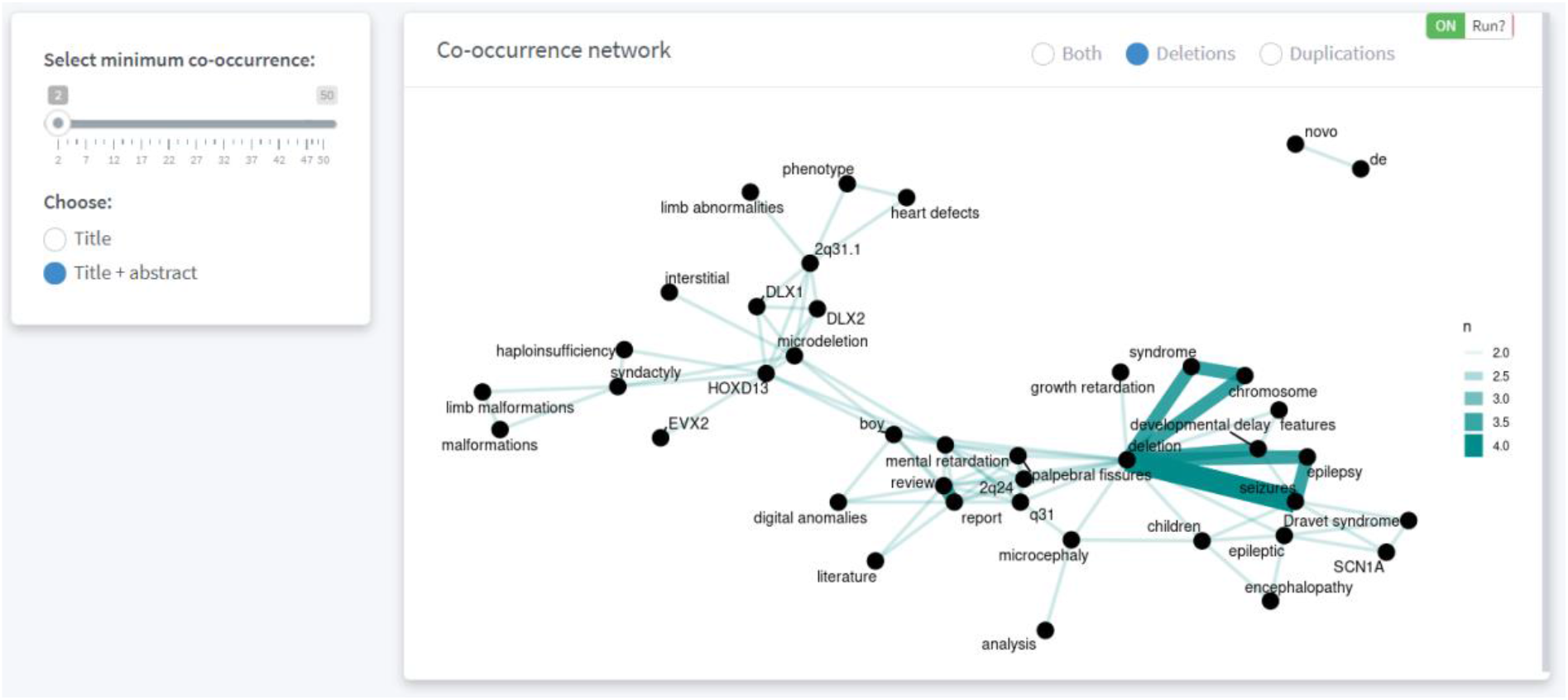
Entity co-occurrence network automatically extracted by CNVxplorer from Pubmed articles associated to the deletion evaluated in Case study 2 of the main text. The figure represents the view generated in the ***Biomedical literature*** tab corresponding to the co-occurrence network of words from articles associated to a deletion in chr2(GRCh37):176065894-176504173. Words are represented as nodes, their co-occurrence across articles is represented as links, and the link thickness is proportional to the number of articles where words co-occurred.

